# The correlation between fetal frontal lobe development parameters and gestational week: A preliminary study

**DOI:** 10.1101/2021.04.20.21255836

**Authors:** Xu Pingping, Zhang Dirong, Shi Yu, Li Dandan, Li Yuan, Kong Fengbei

## Abstract

**Objective:** To establish a reference range of developmental parameters related to the frontal lobe of normal fetuses in different gestational weeks, which can provide a basis for the evaluation of normal development of the frontal lobe and the diagnosis of suspected fetal microcephaly.

**Methods:** A prospective cross-sectional study was performed on 751 normal fetuses between 21 and 36 weeks, and the anterior-posterior diameter and transverse diameter of the frontal lobe were measured in transcerebellar plane. Linear regression analysis was performed by grouping different gestational weeks.

**Results:** Fetal anterior-posterior diameter and transverse diameter of the frontal lobe were positively correlated with gestational week with correlation coefficients 0.871 and 0.971 respectively. There was a linear relationship between gestational weeks and frontal lobe parameters, and the equations were simplified to obtain the simple correlation formula: fetal frontal lobe anterior-posterior diameter(mm)=14.5+0.7×(gestational week-21), frontal lobe transverse diameter (mm)=41+2.5×(gestational week-21).

**Conclusion:** The reference ranges of the frontal lobe anterior-posterior diameter and the frontal lobe transverse diameter of normal fetuses between 21 to 36 weeks in transcerebellar plane were established, which can provide valuable information for the evaluation of normal development of the frontal lobe and the diagnosis of suspicious fetal microcephaly.

**What’s already known about this topic?:** We currently lack a golden standard to diagnosis microcephaly and fetal frontal lobe dysplasia is a clue for prenatal diagnosis of microcephaly.

**What does this study add?:** This study presents a novel evaluation of the fetal frontal lobe to better assess brain development in the transcerebellar plane.

The fetal frontal lobe developmental diameters will be useful to aid diagnosis of suspicious microcephaly and other cortical developmental disorders.

## 1 INTRODUCTION

Microcephaly is often associated with abnormal brain development and may lead to a range of neurological sequelae, such as cerebral palsy, epilepsy, and mental retardation.^1^ There is currently no golden standard for prenatal diagnosis of microcephaly, and a fetal head circumference less than three standard deviations from the mean value of a normal fetus of the same gestational week is a clue for prenatal ultrasound diagnosis of microcephaly,^2^ but the false-positive rate is high.^3^ Microcephaly manifests as a decrease in fetal brain volume, especially in the frontal lobe, and after birth it can manifest as a decrease in head circumference and frontal retraction. Some studies have reported that frontal lobe growth retardation could indicate microcephaly.^4^ A few studies have been reported on the assessment of fetal frontal lobe related developmental parameters in transthalamic plane, but the measurement methods are inconsistent and no uniform reference value is available.^4-7^

The aim of this study was to evaluate the frontal lobe development parameters by using prenatal ultrasound in the transcerebellar plane and investigate the relationship between frontal lobe parameters and gestational week, and establish the normal reference range of frontal lobe anterior-posterior and transverse diameters in different gestational weeks, in order to provide valuable information for assessing the normal development of the frontal lobe and diagnosing fetal microcephaly suspected from 21-to 36-weeks’gestation.

## 2 METHODS

A prospective cross-sectional study was performed on 751 normal fetuses undergoing routine prenatal ultrasound examination between September 2018 and February 2020 in the department of ultrasound of Peking University Shenzhen Hospital. This study was approved by the Peking University Shenzhen Hospital and Health Service Human Research Ethics Committee. The research participants were recruited from pregnant women attending antenatal sonography at our hospital with written informed consent.

### 2.1 Study population

In order to exclude abnormal fetuses and to ensure the relative accuracy of gestational age, the inclusion criteria were as follows: (i) previous regular menstruation with clear date of last menstruation or corrected crown-lump length at early gestational age; (ii) 21 to 36 weeks of gestation; (iii) low risk pregnancy, including the absence of pregnancy risk factors such as diabetes and hypertension; (iv) head circumference confirmed to be within normal range after birth. Exclusion criteria were as follows: (i) known fetal malformations or chromosomal abnormalities; (ii) large discrepancy between the biological gestational week of the fetus and the actual gestational week; (iii) multiple pregnancies.

### 2.2 Ultrasound measurements

GE Voluson E10 and GE Voluson E8 color Doppler ultrasound diagnostics were used with probe frequency 4 to 8 MHz. Firstly, the fetal biparietal diameter, head circumference, abdominal circumference and femur length were measured to assess the biological gestational age of the fetus, and then the fetal structural abnormalities were screened. In the process of detailed screening of fetal central nervous system malformations, fetal frontal anterior-posterior and transverse diameters were measured in transcerebellar plane. Standard transcerebellar plane is defined as follow: transducer is tilted posteriorly to clearly show the brain midline, cavum septum pellucidum, thalamus, cerebellar hemispheres, cerebellar vermis and cisterna magna.^8^ Specific detailed measurements are shown in Figure 1.The frontal lobe anterior-posterior diameter refers to the distance between the junction of the corpus callosum and the anterior border of the cavum septum pellucidum and the medial border of the anterior skull. The frontal lobe transverse diameter refers to the distance from the vertical line made perpendicular to both sides of the skull through the junction of the corpus callosum and the anterior margin of the cavum septum pellucidum. All ultrasound parameters including routine biometrics and frontal lobe diameters were recorded and stored in the research database. Each patient was measured by one experienced sonographer, and each was measured 3 times. When the fetus had poor image acquisition due to positional limitations, the pregnant woman was instructed to perform the scan after moderate activity.

**FIGURE 1.**
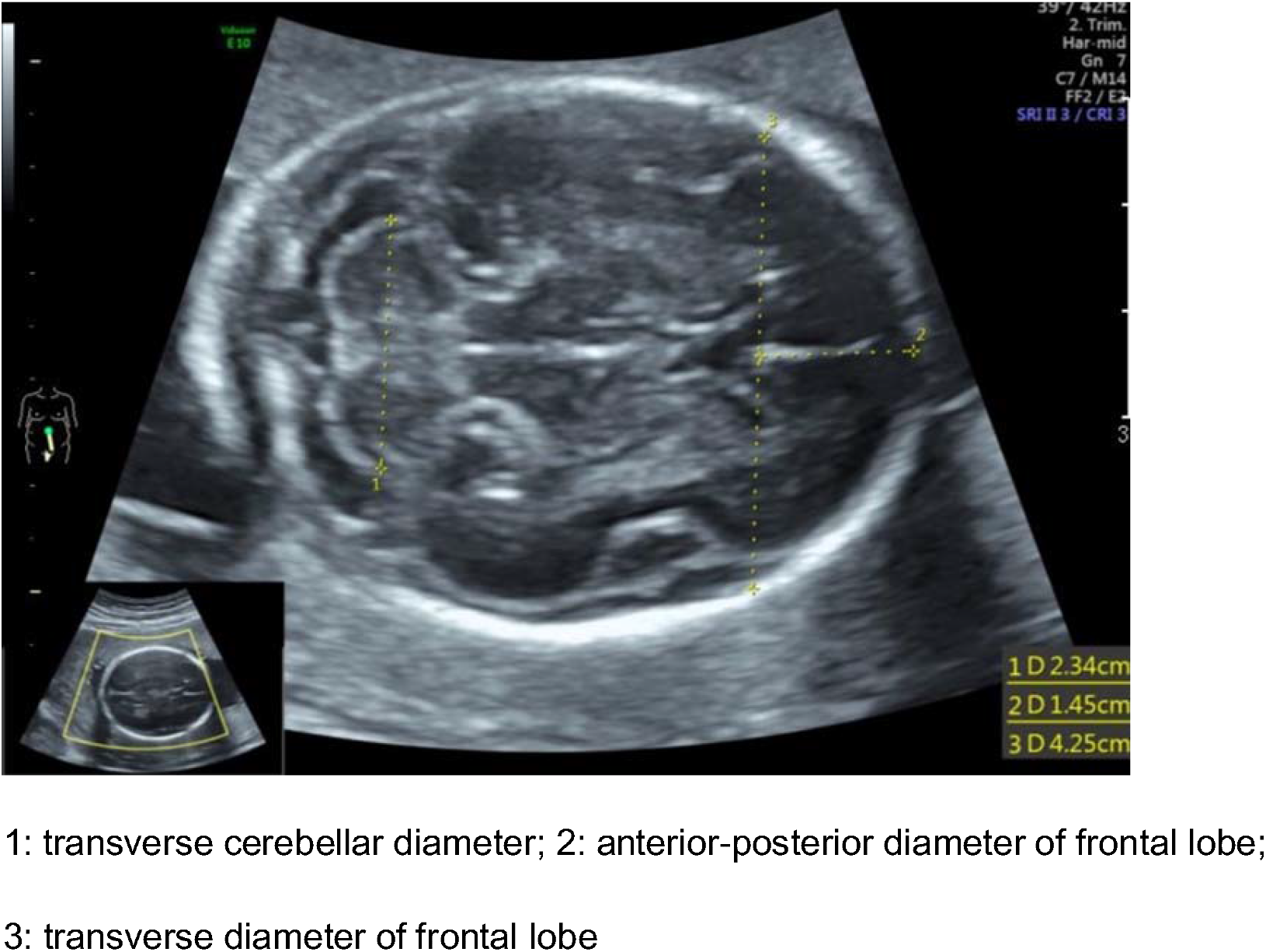
The anterior-posterior diameter and transverse diameter of frontal lobe in transcerebellar plane by two-dimensional ultrasound

### 2.3 Statistical analysis

SPSS 26.0 statistical software was used to process the data. The main processing steps were as follows: (i) Statistical data were collected and divided into 16 groups according to gestational week. The sample size of each group was counted, the mean and standard deviation of each parameter in each group were calculated 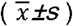; (ii) the normality of each parameter value was tested by the Anderson-Darling test; (iii) for data that conformed to a normal distribution, the 95% reference range was calculated using the two-sided boundary value normal distribution method; for data with a skewed distribution, the normal reference range was calculated using the two-sided boundary value percentile method. The scatter plot of the relationship between the frontal lobe anterior-posterior diameter, transverse diameter of the fetus and the week of gestation was plotted, and a linear regression equation was established by linear regression analysis, with p<0.05 being statistically significant.

## 3 RESULTS

Seven hundred and fifty-one single fetuses with normal fetal sonographic examination of the central nervous system formed our final study sample for analysis, with GAs between 21 and 36 weeks and in mothers aged 20 to 39 years (25.12±2.56 years).

i. The mean, standard deviation and 95% reference range of each parameter at different gestational weeks were calculated, and the bilateral bounds were taken. The data were shown in Table 1.
ii. The scatter plot of the relationship between fetal frontal lobe anterior-posterior diameter and transverse diameter and gestational week were shown in Figures 2 and 3, and the correlation analysis showed that fetal frontal lobe anterior-posterior and transverse diameters were positively correlated with gestational week, with correlation coefficients 0.871 and 0.971 respectively (p values<0.001).
iii. Using gestational week as the independent variable and fetal frontal lobe diameters as the dependent variable in linear regression analysis, the results showed that there was a linear regression relationship between fetal gestational week and frontal lobe diameters. The linear regression equation were as follows: anterior-posterior diameter of frontal lobe(mm)=14.464+0.722× (gestational week-21), transverse diameter of frontal lobe(mm)=40.726+2.611×(gestational week-21).The equation was simplified as fetal frontal lobe anterior-posterior diameter(mm)=14.5+0.7×(gestation week-21) and frontal lobe transverse diameter(mm)=41+2.5×(gestation week-21) respectively.

**TABLE 1.** Fetal anterior-posterior diameter and transverse diameter of frontal lobe at different gestational ages(GA) 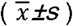 and 95% reference ranges

| GA | Number | Frontal anterior-posterior |  | Frontal transverse |  |
| --- | --- | --- | --- | --- | --- |
|  |  | diameter ( mm ) |  | diameter ( mm ) |  |
| | | ( $\bar{x} \pm s$ ) | 95% reference ranges | ( $\bar{x} \pm s$ ) | 95% reference ranges |
| 21 | 41 | 14.64±1.04 | 12.92~16.74 | 41.45±2.05 | 37.73~44.84 |
| 22 | 79 | 15.10±1.58 | 12.90~18.10 | 43.18±2.22 | 39.61~47.77 |
| 23 | 98 | 15.87±1.56 | 13.20~18.51 | 45.39±2.72 | 40.63~51.40 |
| 24 | 96 | 16.49±1.65 | 13.61~19.26 | 48.01±2.46 | 44.31~52.37 |
| 25 | 45 | 17.37±1.96 | 14.68~21.92 | 51.03±2.89 | 46.30~56.46 |
| 26 | 35 | 18.19±1.63 | 15.24~20.96 | 53.70±2.43 | 49.52~57.66 |
| 27 | 40 | 19.26±1.56 | 16.12~22.19 | 56.34±3.01 | 50.88~62.29 |
| 28 | 40 | 19.97±1.88 | 17.31~23.56 | 59.92±3.11 | 55.97~66.42 |
| 29 | 33 | 20.38±2.36 | 17.06~25.16 | 62.81±3.43 | 57.98~68.45 |
| 30 | 42 | 20.49±2.12 | 16.29~24.04 | 64.52±3.50 | 59.29~70.13 |
| 31 | 44 | 21.77±1.97 | 17.98~25.23 | 68.01±3.31 | 61.69~75.13 |
| 32 | 47 | 22.31±2.00 | 19.62~25.96 | 70.84±3.69 | 66.01~77.93 |
| 33 | 31 | 22.33±1.88 | 18.62~25.72 | 72.98±3.13 | 68.61~79.83 |
| 34 | 28 | 23.64±2.20 | 19.95~27.83 | 73.92±2.54 | 69.45~79.56 |
| 35 | 27 | 24.89±2.02 | 20.60~28.78 | 75.62±2.55 | 71.94~81.90 |
| 36 | 25 | 25.83±1.96 | 21.71~29.83 | 77.91±2.82 | 73.70~83.06 |

**FIGURE 2.**
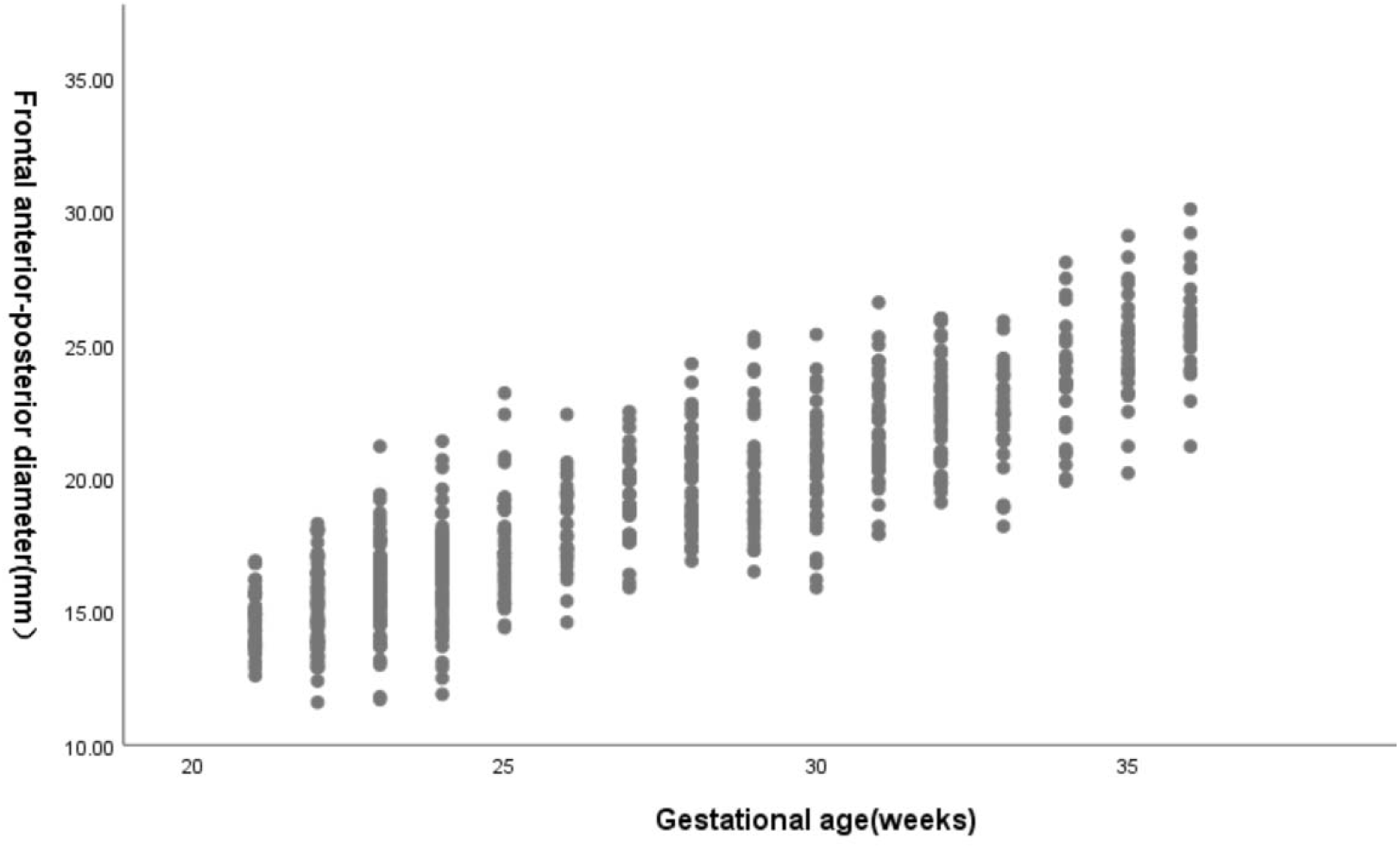
Scatter plot of fetal frontal anterior-posterior diameter with gestational week

**FIGURE 3.**
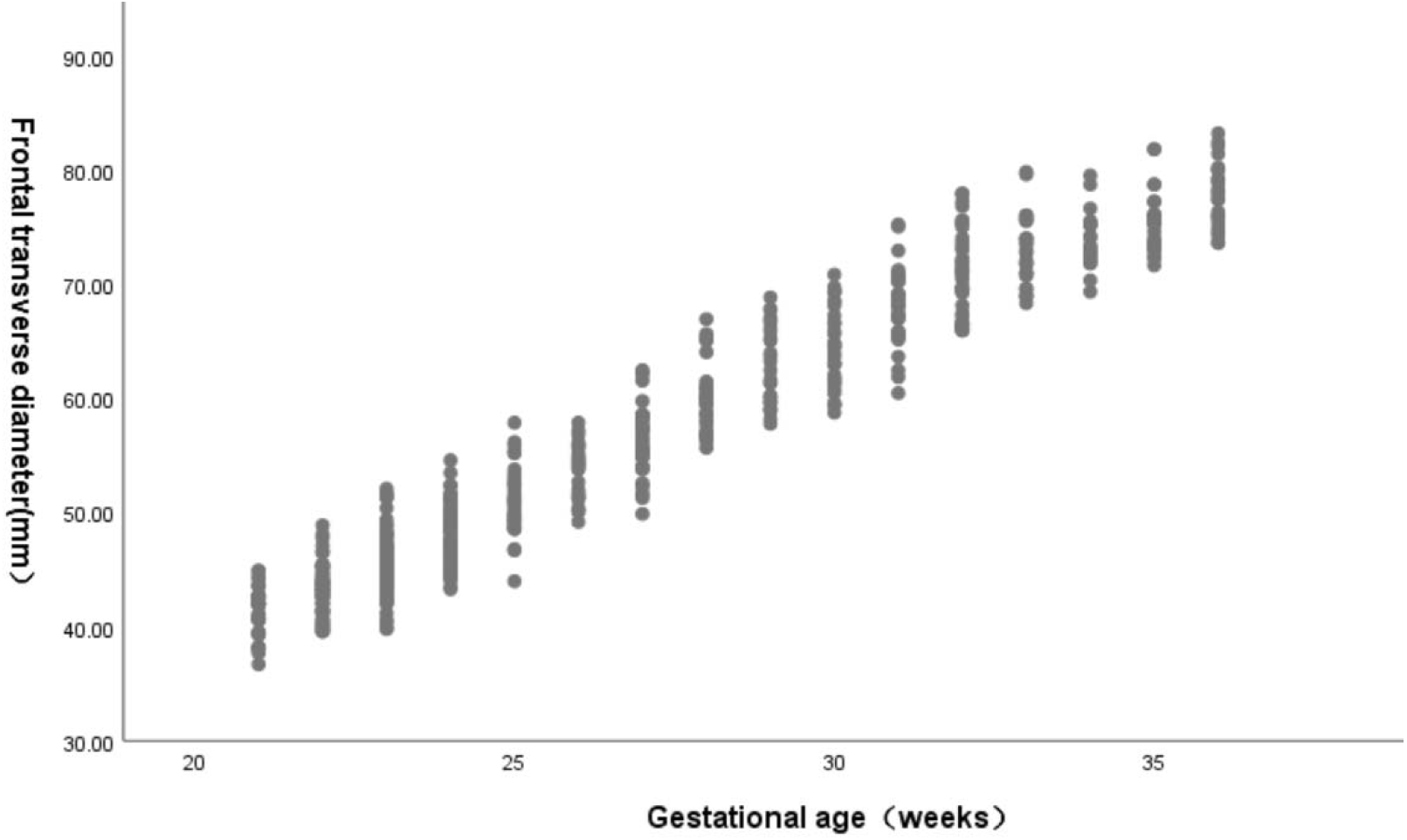
Scatter plot of fetal frontal transverse diameter with gestational week

## 4 Discussion

Microcephaly is a neurodevelopmental disorder, which pathology is mainly manifested as frontal lobe developmental delay. The clinical manifestations are mainly small head circumference especially frontal lobe shrinkage and special facial and intellectual disabilities. There are some causes of fetal microcephaly. Some studies have reported that fetal genetic factors accounted for nearly 50% which the common causes are 21,18,13 trisomy. Fetal intrauterine infection and history of exposure to harmful chemical substances in pregnancy accounted for about 48%, such as intrauterine cytomegalovirus infection and the emergence of Zika virus infection in recent years.^9^ There is currently no golden standard for prenatal diagnosis of microcephaly. Fetal head circumference less than three standard deviations from the mean value of normal fetuses in the same gestational week is the clue for prenatal ultrasound diagnosis of microcephaly, but the false positive rate of microcephaly diagnosis based on this index alone is high up to 43%.^3^ Therefore, when the fetal head circumference is less than three standard deviations from the mean value of normal fetuses in the same gestational week, relevant tests should be performed to exclude chromosomal abnormalities, intrauterine infections and brain developmental delay. Microcephaly is the result of delayed brain development, and frontal lobe involvement is more pronounced than in the mesencephalon and cerebellum, and frontal lobe developmental delay has been reported to be indicative of microcephaly in fetuses.^4,10^

A few studies have been reported on the use of prenatal ultrasound to assess normal fetal frontal lobe development,^4-7^ but these studies select transthalamic plane and the measurements of frontal lobe were inconsistent. In this research, the transcerebellar plane is chosen as the standard measurement view based on the following reasons:(i) the transcerebellar plane is one of the routine sections for screening fetal central nervous system malformations; (ii) the transcerebellar plane is found to show clearer sylvian fissure than the transthalamic plane, and the sylvian fissure is also an important index for evaluating brain development, which is another important index for evaluating brain development and microcephaly in the current study group; (iii)the ratio of transcerebellar diameter to frontal lobe diameter which means caval-calvarial diameter is also an important indicator for the assessment of microcephaly which has already been reported in another study,^11^ but it requires conversion of measurements in the transthalamic and transcerebellar planes, which increases the examination measurement time. In conclusion, our study suggests that the transcerebellar plane is superior to the transthalamic plane in assessing brain development and saves time. In this study, we selected normal fetuses between 21 and 36 weeks of gestation with a successful rate of 95.3%. The linear regression analysis was performed with gestational week as the independent variable and fetal frontal lobe diameters as the dependent variable, and the results showed that there was a linear regression relationship between gestational week and anterior-posterior diameter and transverse diameter, and the linear regression equation was obtained as fetal frontal lobe anterior-posterior diameter(mm)=14.464+0.722×(gestation week-21), frontal lobe transverse diameter(mm)=40.726+2.611 ×(gestation week-21). The equation is simplified as fetal frontal lobe anterior-posterior diameter(mm)=14.5+0.7×(gestation week-21), frontal lobe transverse diameter(mm)=41+2.5×(gestation week-21), which could provide valuable information for the evaluation of fetal frontal lobe development and the suspicious diagnosis of fetal microcephaly by prenatal ultrasound. The results of the present study showed that the correlation coefficient between the frontal lobe transverse diameter and gestational week was higher than frontal lobe anterior-posterior diameter, which was consistent with the result of the study by Brown SA et al.^6^ When fetal head circumference is less than three standard deviations from the mean for fetuses of the same gestational age on prenatal ultrasound, additional tests can be added to assess the developmental parameters of fetal frontal lobe in the transcerebellar plane, and the diagnosis of microcephaly is more likely to be made if the fetal frontal lobe development is more significantly lagging behind.

## 5 Conclusion

The developmental parameters of the fetal frontal lobe were constructed in the transcerebellar plane, and the reference ranges at different gestational weeks were established initially, and the simplified regression equations of the fetal frontal lobe anterior-posterior diameter(mm)=14.5+0.7×(gestation week-21), frontal lobe transverse diameter(mm)=41+2.5×(gestation week-21) were obtained by linear regression analysis. It could provide valuable information for the evaluation of fetal frontal lobe development and the diagnosis of fetal microcephaly, but it needs to be further verified in future work with larger sample sizes and prospective studies.

## Data Availability

Data availability. The full data that supports the findings of this study are available from the corresponding author upon reasonable request.

## ACKNOWLEDGEMENTS

The authors thank all the women who participated in the study and acknowledge their significant contribution. Thank you to Zhang Dirong for helping to perform the ultrasound examinations, Shi Yu for support with statistical analysis, Li Dandan and Li Yuan for data management and collection, and Kong Fengbei for proposing some strategies for writing.

## Notes

**CONFLICT OF INTEREST** The authors declare no potential conflict of interest.

### Competing Interest Statement

The authors have declared no competing interest.

### Funding Statement

Construction of medical key disciplines in Shenzhen city, Grant/Award Number: SZXK051; Shenzhen Virtual Reality Clinical Application Public Service Platform enhancement project, Grant/Award Number: XMHT20190104001; Three Projects Funded Project in Shenzhen city, Grant/Award Number: SZSM201512026

### Author Declarations

This study was approved by the Peking University Shenzhen Hospital and Health Service Human Research Ethics Committee.

## REFERENCES

1. Kaindl AM, Passemard S, Kumar P, et al. Many roads lead to primary autosomal recessive microcephaly. Prog Neurobiol.2010;90(3):363-383.

2. Ashwal S, Michelson D, Plawner L, et al. Practice parameter: evaluation of the child with microcephaly (an evidence-based review): report of the Quality Standards Subcommittee of the American Academy of Neurology and the Practice Committee of the Child Neurology Society.Neurology.2009;73(11):887–897.

3. Chibueze EC, Parsons AJQ, Lopes KDS, et al. Diagnostic accuracy of ultrasound scanning for prenatal microcephaly in the context of Zika virus infection: a systematic review and meta-analysis. Sci Rep.2017;7(1):2310.

4. Goldstein I, Reece EA, Pilu G, et al. Sonographic assessment of the fetal frontal lobe: a potential tool for prenatal diagnosis of microcephaly. Am J Obstet Gynecol.1988;158(5):1057–1062.

5. Caetano AC, Zamarian AC, Araujo Júnior E, et al. Assessment of Intracranial Structure Volumes in Fetuses With Growth Restriction by 3-Dimensional Sonography Using the Extended Imaging Virtual Organ Computer-Aided Analysis Method. J Ultrasound Med.2015;34(8):1397–1405.

6. Brown SA, Hall R, Hund L, et al. A Novel Approach to Prenatal Measurement of the Fetal Frontal Lobe Using Three-Dimensional Sonography. J Reprod Med.2017;62(3-4):119–126.

7. Yaniv G, Katorza E, Tsehmaister Abitbol V, et al. Discrepancy in fetal head biometry between ultrasound and MRI in suspected microcephalic fetuses. Acta Radiol.2017;58(12):1519–1527.

8. Malinger G, Paladini D, Haratz KK, et al. ISUOG Practice Guidelines (updated): sonographic examination of the fetal central nervous system. Part 1: performance of screening examination and indications for targeted neurosonography. Ultrasound Obstet Gynecol.2020;56(3):476–484.

9. Von der Hagen M, Pivarcsi M, Liebe J, et al. Diagnostic approach to microcephaly in childhood: a two-center study and review of the literature. Dev Med Child Neurol.2014;56(8):732–741.

10. Borenstein M, Dagklis T, Csapo B, et al. Brachycephaly and frontal lobe hypoplasia in fetuses with trisomy 21 at 11+0 to 13+6weeks. Ultrasound Obstet Gynecol.2006;28(7):870–875.

11. Persutte WH, Coury A, Hobbins JC. Correlation of fetal frontal lobe and transcerebellar diameter measurements: the utility of a new prenatal sonographic technique. Ultrasound Obstet Gynecol.1997;10(2):94–97.

